# Protocol of the COVID-19 Health Adherence Research in Scotland Vaccination (CHARIS-V) study: Understanding the influence of vaccination decisions on adherence to transmission-reducing behaviours in a prospective longitudinal study of the Scottish Population

**DOI:** 10.1101/2021.05.03.21256503

**Authors:** Marina Maciver, Chantal Den Daas, Marie Johnston, Diane Dixon, Gill Hubbard, on behalf of the CHARIS Consortium

## Abstract

**Introduction:** The global population has been asked to live under tight restrictions to slow the spread of SARS-CoV-2. Transmission-reducing behaviours (TRBs), (physical distancing, hand washing, wearing a face covering) were introduced by governments in 2020 prior to vaccine availability. People should maintain TRBs throughout the vaccination programme, because there is much uncertainty about the vaccine efficacy, immunity duration, whether there will be the requirement for booster vaccines, and whether vaccinated individuals can be carriers of the virus. This study will explore the effect of the vaccination programme in Scotland on adherence to TRBs.

**Methods and analysis:** Telephone interviews will be conducted with participants from the nationally representative CHARIS project that agreed to be contacted for further research. Approximately 200, ten-minute telephone interviews will be conducted. Data will be collected on vaccine uptake or intention and adherence to TRBs. Univariate and multivariate logistic regression analyses and moderation analyses will be used to analyse the data collected. Ethical approval was granted by the Life Sciences and Medicine School Ethics Review Board (SERB) at the University of Aberdeen.

**Discussion:** CHARIS-V will provide an understanding of the effect of the vaccination programme on adherence to TRBs in Scotland. Findings should be useful to governments and public health agencies for the current COVID-19 pandemic and vaccination programme and assist in the management of any future outbreaks.

## INTRODUCTION

A new coronavirus (SARS-CoV-2) emerged in Wuhan, China in December 2019 leading to an acute respiratory syndrome (COVID-19) in humans and the initiation of a worldwide pandemic [1]. The UK has one of the highest numbers of recorded cases and deaths from COVID-19 in Europe [2]. In order to suppress the virus to the lowest possible level, vaccination programmes have been implemented [3]. Stringent transmission-reducing behaviours (TRBs) are also in place across the UK to slow the spread of SARS-CoV-2 [4]. Although there is now a national vaccination programme, uncertainties currently remain as to whether people who have had the vaccine are fully protected, how long immunity lasts, whether there will be the requirement for repeated booster vaccines and whether vaccinated individuals can transmit the virus to others [5]. As there are so many uncertainties surrounding COVID-19 vaccination efficacy, understanding the factors determining adherence to TRBs is important.

### Transmission-reducing behaviours

In Scotland, guidelines on TRBs were introduced by the Scottish Government in their strategic framework [8]. It has been necessary for the nation to stay at home for much of 2020 and the beginning of 2021, only going outside for a few essential reasons. When going outside, people have been required to adhere to strict physical distancing, hand-washing measures and the wearing of face-masks [8]. Due to the mechanism of spread of COVID-19, via respiratory droplets,[14] TRBs should be effective at slowing the spread of the virus. A systematic review and meta-analysis by Chu et al. [15] showed that transmission of viruses was reduced if physical distancing measures were adhered to. Transmission was less if a distance of at least one metre was maintained and as distance increased transmission rates decreased.

COVID-19 is also thought to be spread by coming into contact with surfaces infected by respiratory droplets and subsequently touching the face [16]. Touching the face if frequent and habitual [18]. Since people cannot be prevented from touching their face, then hand-hygiene is considered important in reducing the transmission of the virus [17]. The UK government has promoted good hand-hygiene as the washing of your hands frequently and thoroughly, with soap and water for a minimum of 20 seconds [19,20]. Hand washing has been demonstrated to reduce the occurrence of infectious diseases [21-23]. Another important TRB is the wearing of a face-covering in public places. The World Health Organisation (WHO) recommend that a face-covering is worn by members of the public at times where physical-distancing is difficult [24]. It is anticipated that restrictions will be able to be relaxed as the nation gets vaccinated. However, it remains important at present, and during the roll-out of the vaccination programme that individuals adhere to these TRBs [6,7,9,10].

It is currently unclear what effect the COVID-19 vaccination programme will have on adherence to TRBs. A major concern is that one unintentional and undesirable effect will be that once individuals have been vaccinated then they will no longer adhere to TRBs [12]. Indeed, an online survey of 1,706 adults conducted a week prior to the first vaccinations being carried out in the UK (8^th^ December 2020) found that 50% of participants reported that after having the vaccine they would ‘probably still follow whatever coronavirus rules or restrictions were in place as strictly as they had before getting a vaccine’, 29% said that they would ‘probably stick to them less strictly’ and 11% said that they would ‘probably no longer follow the rules or restrictions’ [25]. Therefore, there is an urgent need to understand people’s adherence to TRBs in relation to vaccine behaviour and beliefs about the vaccine.

### Theoretical Approaches to Human Motivation and Behaviour Explaining Adherence to TRBs

In a pandemic, it is useful to be able to understand how people respond to the threat of illness and their reaction to recommendations or mandated restrictions in relation to behavioural changes. We aim to do this by exploring adherence to TRBs through the lens of 3 theoretical frameworks, namely the common - sense model of self - regulation, social cognitive theory and protection motivation theory. The common-sense model of self-regulation (CS-SRM) is a theoretical framework that explains how individuals respond to and manage to health threats, in this instance COVID-19 [41].CS-SRM identifies cognitive and emotional illness representations and their affect on behavioural responses. Cognitive representations include identity (diagnosis label, symptoms), what caused the illness, time-line of the illness, personal consequences of the illness and curability or controllability of the illness by the individual or medical treatment [42]. Emotional representations are the emotions (e.g. anxiety or stress) illness threat gives rise to. An individual’s way of coping with a threat may be cognitive or emotional, adherence to TRBs may be to reduce anxiety or control disease exposure.

Social Cognitive Theory (SCT) describes the influences of personal experiences, others actions and environmental factors on health behaviours [43]. Self-efficacy and outcome expectancies are key constructs of SCT. Self-efficacy is the belief that an individual is capable of carrying out a recommended behaviour and achieving a desired outcome (ie. not catching COVID-19) and outcome expectancies refer to the belief that carrying out a behaviour will give them the desired outcome (eg. receiving the COVID-19 vaccination will reduce my risk of being seriously ill from COVID-19) [44].

Protection Motivation Theory (PMT) [26,27] is a psychological theory concerned with how individuals perceive and respond to a threat. Several studies of COVID-19 transmission-reducing behaviours have suggested, using Protection Motivation Theory, that adherence to TRBs is influenced by both threat appraisal and coping appraisal [28-30]. Perceived severity of the illness and perceived vulnerability to the disease are the factors considered in the threat appraisal; the coping appraisal is concerned with response efficacy, i.e., the perceived ability to carry out the behaviour required to ward off the threat [31,32]. Using this theory, individuals who believe: a) that they would become seriously ill if they caught COVID-19, b) that it is likely that they could catch SARS-CoV-2, c) physical distancing, hand washing and wearing a face covering will reduce the risk of getting COVID-19, and d) that they are able to adhere to these risk-reducing behaviours are more likely to adhere to these transmission-reducing behaviours than individuals who do not have these beliefs.

The COVID-19 Health Adherence Research In Scotland (CHARIS) project found that perceived severity and response efficacy, self-efficacy and intention and timeline and cause were predictive in univariate and multivariate analyses of physical distancing, hand washing and face covering [33]. However, people’s beliefs could change after they receive the vaccine. Vaccination may provide people with a sense of security and therefore change their perceived vulnerability to the disease [12].

### CHARIS-V STUDY, AIMS AND OBJECTIVES

CHARIS-V, is an extension of the CHARIS project, and aims to investigate adherence to physical distancing, hand-washing behaviour and the wearing of face coverings during the implementation of the COVID-19 vaccination programme and explain variation in adherence to TRBs.

The following research questions will be addressed:

1. How many people have been, or intend to get vaccinated?
2. Does adherence to TRBs (physical distancing, face covering, hand washing) prior to the vaccination programme predict vaccination behaviour/intention?
3. Do behavioural beliefs prior to the vaccination programme predict vaccination behaviour/intention?
4. To what extent does vaccination response efficacy predict vaccination behaviour/intention?
5. Does vaccination behaviour/intention predict adherence to TRBs during the vaccination programme?
6. Do sociodemographic variables moderate associations between vaccination behaviour/intention and adherence to TRBs during the vaccination programme?

## METHODS

### Design and Participants

The CHARIS project was a serial, weekly, nationally representative, cross-sectional study of randomly selected adults in Scotland [34]. In total, 4529 participants from the CHARIS project had consented to be approached by the research team with regards to participating in subsequent CHARIS project studies. For the CHARIS-V study, participants from the CHARIS project that consented to be contacted for follow-up will be contacted by telephone for the purposes of recruiting them to the CHARIS-V study. Again, interviews will be carried out by telephone, so participants do not require financial means or computer literacy to take part. CHARIS-V is a prospective longitudinal study.

### Statistical Power

Using G-Power, assuming alpha of .05, 80% power, effect size 0.15 and 22 predictors will require a sample size, N=163 people. To allow the potential for exploration of subgroups (age, gender, SIMD) a sample size of N= 200 will be recruited.

### Patient and Public Involvement Statement

The CHARIS project questionnaire was reviewed by two Patient and Public Involvement (PPI) groups [34].The CHARIS-V study protocol was discussed with one of the PPI groups involved in the development of the CHARIS project questionnaire (National Research Scotland Primary Care PPI group), who considered the study to potentially have important data to inform public health policies during the pandemic. This group was pivotal in identifying the topic of vaccination and TRBs as a priority during the design of CHARIS-V and its questionnaire.

### Measures

#### COVID-19 Vaccination intention and behaviour

Participants will be asked if they have been invited to get the Covid-19 vaccine and can respond with yes or no. If they have been invited, they will be subsequently asked if they had already had the vaccine (yes, no). If they had already had the vaccine then they will be asked how many doses (one, two). Those that report that they had had one dose will be asked if they intend to get the second dose when they are invited (yes definitely, yes probably, no definitely, no probably). Participants will be asked how long ago they had their last Covid-19 vaccination (within the last week, 2 to 3 weeks ago, longer than 3 weeks ago). If participants had not already had the vaccine (i.e. had not yet been invited to receive the vaccine, or had been invited but not yet had it) then they will be asked if they intend to get the vaccine (yes definitely, yes probably, no definitely, no probably).

#### COVID-19 Vaccination response efficacy

Response efficacy is a key concept of PMT [26], an important theoretical framework for understanding COVID-19. Two questions will be used to determine Covid-19 vaccination response efficacy in CHARIS-V: How much, if at all, do you agree or disagree with the following statements: 1) People who have the vaccine are less at risk of getting Covid-19 than people who have not had the vaccine, 2) People who have the vaccine are less likely to spread the coronavirus than people who have not had the vaccine. Participants will indicate the extent to which they agreed or disagreed using a 5-point response scale (strongly agree, tend to agree, tend to disagree, strongly agree) and can also respond as don’t know or prefer not to say.

#### Transmission-reducing behaviours

Adherence to TRBs will be assessed with three items for the three domains of behaviour, physical distancing, handwashing and wearing a face-covering [8, 35]. In CHARIS-V, adherence to transmission-reducing behaviours during vaccination was measured among participants who answered ‘yes’ to the following question: ‘Did you go outside your home last week?’ using the following three items: ‘Please tell me, how often, if at all, in the past week, you…1) stayed 2 metres (6 feet) away from other people, except those who live in your household, 2) washed your hands as soon as you got home, 3) wore a face covering when you were in a shop.’ A 5-point response scale (always, most times, sometimes, rarely, never) will be used to enable participants to indicate the extent to which they have adhered to each behaviour. For individuals who have indicated that they have been vaccinated the following question will be asked: ‘Now that you have been vaccinated, will you be more or equally likely or less likely to keep 2 metres distance from other people not in your household? A 3 point response scale (more likely, equally likely, less likely) will be used to measure their behaviour. For participants who have not yet been vaccinated but who have indicated that they intend to be vaccinated the following question will be included: ‘When you have been vaccinated, compared to what you do now, will you be more or equally likely or less likely to keep 2 metres distance from other people not in your household? A 3 point response scale (more likely, equally likely, less likely) will be used to measure their anticipated behaviour.

#### Sociodemographic variables

Sociodemographic information was collected during the initial CHARIS project [34]. Information collected included: location, Scottish Index of Multiple Deprivation (SIMD), age, gender, ethnicity, number of adults and children living in the household, household tenure and employment status [36].

#### Theory-based variables

The CHARIS project’s measures of beliefs from three theories will be used in CHARIS-V: beliefs about risks (Protection Motivation Theory); beliefs about illness (Common-sense Self-Regulation Model), and beliefs about TRBs (Reasoned Action Approach).

#### Data Processing and Analysis

Data will be stored on a secure University server. An individual’s data will include a subject identification number, this will be used to link the data from CHARIS-V to the CHARIS project. Identifying information used to conduct the telephone interviews will be securely stored in a separate file not used in the data analysis. Individuals will be asked to provide consent for the linkage of the data.

The data will be analysed using SPSS 25.0. Answers ‘I don’t know’ and ‘I prefer not to say’ will be treated as missing values for all variables and will be excluded from the analyses. Summary statistics, including means, standard deviations, medians and ranges for continuous variables and frequency distributions for binary and categorical variables will be used (RQ1). The Pearson bivariate correlation will be used to examine any relationships between variables. P values of < 0.05 will be treated as being statistically significant.

Univariate and multivariate logistic regression analysis will be used to assess whether adherence to each of the TRBs prior to vaccination (RQ2), theoretical factors (RQ3), vaccine response efficacy (RQ4) are predictive of vaccination behaviour/intention. In addition, we will assess whether vaccination behaviour or intention is associated with adherence in 2021 (RQ5). A binary outcome of non-adherence, defined as individuals who sometimes, rarely or never adhere to TRBs, compared to participants who always or mostly adhere will be used. Multivariate models will be constructed using the entry method, including those variables that are found to be significantly associated in the univariate analysis.

Moderation model analyses will be used for the analysis of the relationship between vaccination behaviour, sociodemographic characteristics and theoretical factors. Hayes’ PROCESS macro, model 1 [40] will be used in SPSS. For the moderation analysis (RQ6), vaccine behaviour will be entered into a regression model with one of the moderator variables (sociodemographic variable, threat perception, behavioural beliefs) and the outcome of adherence. The interaction term between vaccination behaviour and the moderator variable will be entered in the next step. A 95% bootstrap confidence interval (CI) method will be used for the analyses. Simple slopes for the association between vaccination behaviour and adherence will be plotted for different levels of the sociodemographic variables and theoretical factors, for the continuous variable we will plot low (−1 SD below the mean), moderate (mean), and high (+1 SD above the mean) levels [45].

#### Ethics

Ethical Approval for the CHARIS project and the subsequent CHARIS-V study was granted by the Life Sciences and Medicine School Ethics Review Board (SERB) at the University of Aberdeen. Consent will be sought from each participant for their participation in CHARIS-V and for linkage of CHARIS-V data with the data from the CHARIS project. Participants can opt to not give an answer to any of the questions if they so wish.

## DISCUSSION

The CHARIS-V study will provide an understanding of the effect of the vaccination programme on adherence to TRBs. By using data from the first CHARIS project collected prior to the implementation of the vaccination programme with the data collected during CHARIS-V at the start of national roll-out of the vaccination programme in Scotland, the study will be able to predict adherence to TRBs over time, whether vaccine intention/behaviour is predictive of adherence to TRBs, whether previous adherence to TRBs is predictive of vaccine behaviour/intention, and whether sociodemographic factors are predictive of vaccine intention and adherence to TRBs. The study can be used to tailor and target public health interventions and is likely to make a unique contribution to the emerging body of international work about TRBs during pandemics and during a vaccination programme.

### Strengths

The CHARIS-V study has several strengths. It will be one of the first studies to provide useful information on adherence to TRBs and the influence of vaccination on these behaviours during the commencement of the vaccination programme. CHARIS-V is a prospective longitudinal study and will be conducted during the vaccination programme allowing for real-time information to inform public health policies in this current pandemic and in future outbreaks requiring vaccination. The timing of CHARIS-V study places it in a unique position to assess potentially changing adherence to TRBs as people are vaccinated. The CHARIS-V study data is collected via telephone, requiring the questions to be easily understood. This reduces significant barriers to people participating in the study, allowing more vulnerable populations to be included [37,38]. Recruitment to the study will be from the main CHARIS study participants and aims to be a representative sample, making the findings generalisable to the Scottish population.

### Limitations

A limitation of this study is that we require individuals to self-report due to using telephone interviews to collect data. Measuring behaviour by self-report may cause the results to be subject to social desirability bias. However, it would be impossible to measure adherence to all the TRBs included in CHARIS-V by any other method. In addition, observational measures for adherence to TRBs could not be linked to individual vaccination behaviour. We believe the advantages provided by telephone interviews, such as population reach and generalisability outweigh the disadvantages. Another limitation in this study is that not everyone in Scotland will be invited to be vaccinated at the same time potentially resulting in variation in assessing behaviour and intention. Although this is a potential limitation of this study, variation in behaviour and intention is expected to be low as vaccine uptake in Scotland is expected to be high [8]. In addition, it will give information on adherence to TRBs at a time when only a small proportion of the population have been vaccinated, and adherence to TRBs are most crucial.

## CONCLUSION

CHARIS-V is likely to make a significant contribution to the evidence about adherence to TRBs and the influence vaccine intention and behaviour has on these behaviours. Findings should be useful to governments and public health agencies for the current COVID-19 pandemic and vaccination programme and assist in the management of any future outbreaks.

## Data Availability

Data availability not applicable to this article

